# MYD88 L265P mutation in primary central nervous system lymphoma is associated with better survival: A single centre experience

**DOI:** 10.1101/2020.09.06.20185827

**Authors:** Olimpia E. Curran, Michael T. C. Poon, Louise Gilroy, Antonia Torgersen, Colin Smith, Wael Al-Qsous

## Abstract

**Background:** The *Myeloid differentiation primary response gene* (*MYD88*) mutation in primary central nervous system lymphomas (PCNSL) may be associated with unfavourable prognosis, however the evidence remains limited. We aimed to comprehensively characterise PCNSLs by integration of clinicopathological, molecular, treatment and survival data.

**Methods:** We retrospectively identified and validated 57 consecutive patients with PCNSLs according to the 2017 WHO classification of lymphoid neoplasms over a 13-year period. Formalin-fixed paraffin-embedded tumour samples underwent real-time allele-specific polymerase chain reaction assay to detect *MYD88* mutation. We used multivariable Cox regression for survival analysis including age, treatment, and *MYD88* as covariates. We searched the literature for studies reporting demographics, treatment, *MYD88* and survival of PCNSL patients, and incorporated individual-patient data into our analyses.

**Results:** The median age was 66 years and 56% were women. All 57 patients had non-germinal PCNSL and the majority (81%) received either single or combined therapies. There were 46 deaths observed over the median follow-up of 10 months. *MYD88* mutation status was available in 41 patients of which 36 (88%) were mutated. There was an association between *MYD88* mutation and better survival in the multivariable model (hazard ratio [HR] 0.34; 95% confidence interval [CI] 0.12-0.95; p=0.039) but not in a univariable model. After incorporating additional 18 patients from the literature, this association was reproducible (HR 0.31, 95% CI 0.13-0.77, p=0.012).

**Conclusions:** Adjusting for confounders, *MYD88* mutation is associated with better survival. While further validation is warranted, identification of *MYD88* mutation can identify patients who may benefit from novel targeted therapies.

**Key points:**

1. *MYD88* mutation is common in PCNSLs.
2. *MYD88* mutation in PCNSLs is associated with better survival after adjusting for age at diagnosis and treatment.
3. Identification of *MYD88* mutation in PCNSLs can identify patients who may benefit from novel targeted therapies and enhance survival.

**Importance of the study:** PCNSLs are rare and associated with lower survival than their systemic counterparts. The emergence of new molecular targets in PCNSLs, such as mutations in the *MYD88* gene, offers hope for more effective therapeutics. Few studies have investigated the association between *MYD88* mutation and survival. These studies, however, are limited by inconsistent inclusion of clinical variables and suboptimal analytic approach, such as overfitting model or incomplete adjustment for important confounders. Our study integrates treatment, molecular and survival data for 57 patients diagnosed with PCNSL. We demonstrate that without adequate adjustment for confounders such as age at diagnosis and treatment, *MYD88* mutation does not affect survival. However, a multivariable survival model including these variables shows *MYD88* mutation to be associated with better survival. While further validation of this association is warranted, our findings suggest that identification of *MYD88* mutation can identify patients who may benefit from novel targeted therapies and enhance survival.

## INTRODUCTION

Malignancies of lymphoid origin in the brain are rare and associated with poor survival ^1^. The most common diagnostic entity is a PCNSL, defined as diffuse large B-cell lymphoma (DLBCL) arising within the brain, spinal cord, leptomeninges or eye with no evidence of disease elsewhere ^2,3^. The majority of cases exhibit a non-germinal centre cell origin by immunohistochemistry (IHC), corresponding to late germinal centre exit B cells with blocked terminal B-cell differentiation ^3,4^. Brain biopsy is considered the gold standard for PCNSL diagnosis ^5^. Treatment options include single therapies, such as chemotherapy or radiotherapy, or a combination of both, or with an autologous stem-cell transplantation (ASCT) ^5^. Chemotherapy options are typically adapted from systemic regimens, which are compromised by their limited ability to cross the brain-blood barrier ^6^. The emergence of new therapies designed for molecular targets, such as *MYD88* signalling pathway ^7^, may offer additional treatment options for patients affected by these rare tumours.

The *MYD88* L265P mutation is frequent in PCNSLs and has recently been identified as a potential diagnostic marker ^8–12^. The *MYD88* gene codes for a B-cell signalling adaptor protein ^13^. A switch of amino acid leucine to proline at position 265 (L265P) leads to constitutive activation of the nuclear transcription factor kappa B (NF-κB) signalling. This pathway is frequently dysregulated in systemic DLBCLs ^7,10^ and *MYD88* signalling pathway has been evaluated as a potential therapy for DLBCLs. Ibrutinib, a selective inhibitor of Bruton tyrosine kinase (BTK), has also been successfully tried in systemic and CNS DLBCLs with *MYD88* mutation ^14–17^.

The prognostic value of *MYD88* L265P mutation in PCNSLs remains inconclusive. Several studies reported no effect ^9,18–20^ or unfavourable outcome on overall survival in PCNSLs ^21,22^. A recent meta-analysis of *MYD88* mutation in DLBCLs from any site has showed no impact on overall survival ^10^.

In this study, we reviewed 57 PCNSLs diagnosed at a single neuropathology centre. We retrospectively tested formalin-fixed paraffin-embedded (FFPE) brain tissue for the presence of *MYD88* L265P mutation and assessed its associations with overall survival taking clinicopathological features and treatment regimens into account. We validated our findings in a larger cohort of PCNSLs identified through a systematic literature search.

## MATERIALS and METHODS

### Patients

Histological sections of 57 PCNSLs diagnosed at our centre between 1 January 2007 and 1 March 2020 were retrieved from the archives. Clinical characteristics collected from the neuropathological reports included sex (female or male), age (<60, 60-69 or 70+ years), location of lesion (deep or superficial), and tumour extent (unilateral or bilateral). Information about tumour location and extent were confirmed with imaging reports. We searched local clinical databases for information about received treatment regimes, follow-up and survival. The ethical approval for this study was waived by the Tissue Governance committee of the South East Scotland SAHSC BioResource.

### Histopathology and immunohistochemistry

Haematoxylin and eosin-stained sections were reviewed in order to confirm diagnosis in accordance with the current 2017 WHO classification of lymphoid neoplasms ^23^. Archived immunohistochemistry stained sections were reviewed for expression of B-cell markers, including antibodies against CD20, BCL2, BCL6, MUM1 and CD10. Using the Hans algorithm, cases were further subclassified into germinal B-cell centre (GCB) and non-GCB subtypes^3,24^.

### Sample preparation for *MYD88* mutation analysis

Genomic DNA was extracted from FFPE tissue blocks and analysed using a real-time, allele specific polymerase chain reaction (PCR) analysis. Briefly, DNA was extracted from the FFPE sample using the QIAamp DNA FFPE tissue kit (Qiagen). Real time allele-specific PCR was performed according to Jimenez et al ^25^. The assay carries a limit of detection of 1% when a minimum of 25 ng DNA is utilised.

### Data pooling

A literature search was performed up to May 28, 2020 for published articles in English using PubMed and Embase. The searching details were *MYD88* and primary central nervous system lymphoma. The search was limited to human studies. We excluded patients with systemic DLBCLs, primary vitreoretinal lymphoma and immunocompromised cases, either HIV-or EBV-positive as all these entities show distinct clinicopathologic features ^26^. We also excluded studies on liquid biopsies. We searched for detailed information about treatment and survival in adult PCNSL patients who were tested for the presence of *MYD88* L265P mutation using molecular techniques. Studies reporting *MYD88* expression using IHC were not included, unless validated with molecular techniques.

### Statistical analysis

Overall survival (OS) was defined as the time from the date of surgery to the date of death or with censoring on the date of last available follow up. Survival curves were estimated using the Kaplan–Meier method and compared using log-rank test. We used univariable and multivariable Cox regression to assess the effect of *MYD88* mutation and other clinical predictors on overall survival. The association between *MYD88* mutation and survival was the main effect in the multivariable Cox regression with age and treatment modality as covariates. We chose these covariates because these are the strongest confounders and we did not include any further putative confounders to avoid overfitting. All survival analyses and graphs were produced with R statistical software (R version 4.0.0) using *tidyverse*, *survminer*, and *survival* packages.

## RESULTS

### Clinical data

In the period from 1 January 2007 and 1 March 2020 a total of 57 PCNSL patients were diagnosed at our neuropathology centre. The median patient age at the time of biopsy was 66 years (range: 31–78 years). Females constituted 56% of patients (female/male ratio 1.3:1). Nineteen (33%) and 38 patients (67%) had a superficial lesion and a deep lesion, respectively. Unilateral involvement was recorded in 42 (74%) patients and bilateral in 15 cases (26%). In all cases, the diagnosis was made on brain tissue biopsy. Following surgery, 25 (44%) patients received a single, and 21 (37%) a combination therapy. Chemotherapy combined with radiotherapy, chemotherapy alone or radiotherapy alone were given to 18 (32%), 17 (30%) and 8 (14%) patients, respectively. Three patients (5%) received autologous stem cell transplant following chemotherapy. Eleven patients (19%) were not fit and did not receive any treatment after the diagnostic biopsy. Details of individual patient treatments are shown in Supplementary Table S1. The 1-year, 3-year and 5-year survival based on Kaplan-Meier estimation were 52% (95% confidence interval [CI]: 40.5-67.6%), 22.5% (95%CI: 13.1-38.8%) and 7.5% (95%CI: 2.6-21.9%), respectively. At the last follow up, 46 patients had died, and 11 patients were alive. The median follow-up time was 10 months (range: 0-96 months).

### IHC and molecular data

All our PCNSLs were DLBCL of non-GCB subtype. *MYC* analysis by fluorescent in-situ hybridisation (FISH) was available for 23 cases (15 *MYD88* mutant, 2 *MYD88* wild-type and 6 *MYD88* mutation status undetermined) none of which showed evidence of a *MYC* rearrangement.

### Data pooling

Seven studies fulfilled our literature search criteria. Among these studies, a study of Yamada et al.^9^ provided individual patient data on treatment, *MYD88* mutation status and overall survival for additional 18 PCNSL cases.

### *MYD88* L265P mutation analysis

Tissue blocks were not available for 3 (5%) cases. *MYD88* mutation analysis was performed in 54/57 specimens. In 13 (23%) of the cases the DNA content was too low for reliable testing. There were no major differences in clinical characteristics between the patients with missing *MYD88* data (16/57) and those included in the subsequent survival analyses (41/57) apart from treatment variable (Supplementary Table S2). Overall, good quality genomic DNA was available for 41 (72%) cases. The *MYD88* c.794T>C substitution status was detected (mutant) in 36/41 (87.8%) patients. There were no significant differences between mutant and wildtype *MYD88* patients (Table 1). There was a tendency for *MYD88*-wildtype cases to have a shorter follow-up, but this difference was not statistically significant (2 vs 14 months, p=0.075). The same analysis performed with additional 18 PCNSL cases from Yamada *et al*. ^9^ showed no significant differences between mutant and wildtype *MYD88* patients. However, the median follow-up was significantly shorter for wild type patients in comparison to mutated patients (3 vs 15 months, p=0.045) (Table 2).

**Table 1.** Details of 41 Scottish PCNSL patients with known MYD88 L265P mutation status.

| Characteristic | Wild type, N = 5<sup>1</sup> | Mutated, N = 36<sup>1</sup> | p-value<sup>2</sup> |
| --- | --- | --- | --- |
| Age group |  |  | >0.9 |
| <60 years | 1 (20%) | 11 (31%) |  |
| 60-69 years | 2 (40%) | 14 (39%) |  |
| 70+ | 2 (40%) | 11 (31%) |  |
| Sex |  |  | 0.4 |
| Female | 4 (80%) | 19 (53%) |  |
| Male | 1 (20%) | 17 (47%) |  |
| Extent |  |  | 0.6 |
| Bilateral | 2 (40%) | 8 (22%) |  |
| Unilateral | 3 (60%) | 28 (78%) |  |
| Deep location | 4 (80%) | 22 (61%) | 0.7 |
| Treatment |  |  | 0.3 |
| None | 2 (40%) | 6 (17%) |  |
| Single | 2 (40%) | 12 (33%) |  |
| Combined | 1 (20%) | 18 (50%) |  |
| Median follow-up (months) | 2 (0, 4) | 14 (4, 29) | 0.075 |

<sup>1</sup> Statistics presented: n (%); median (IQR)
<sup>2</sup> Statistical tests performed: Fisher's exact test; chi-square test of independence; Wilcoxon rank-sum test

**Table 2.** Details of pooled data from 41 Scottish PCNSL patients and additional 18 PCNSL cases from Yamada *et al*. 2015 with known MYD88 L265P mutation status, N=59.

| Characteristic | Wild type, N = 6<sup>7</sup> | Mutated, N = 53<sup>7</sup> | p-value<sup>2</sup> |
| --- | --- | --- | --- |
| Age group |  |  | >0.9 |
| <60 years | 2 (33%) | 16 (30%) |  |
| 60-69 years | 2 (33%) | 22 (42%) |  |
| 70+ | 2 (33%) | 15 (28%) |  |
| Sex |  |  | 0.7 |
| Female | 4 (67%) | 26 (49%) |  |
| Male | 2 (33%) | 27 (51%) |  |
| Extent |  |  | 0.6 |
| Bilateral | 2 (40%) | 8 (22%) |  |
| Unilateral | 3 (60%) | 28 (78%) |  |
| Unknown | 1 | 17 |  |
| Location |  |  | 0.7 |
| Superficial | 1 (20%) | 14 (39%) |  |
| Deep | 4 (80%) | 22 (61%) |  |
| Unknown | 1 | 17 |  |
| Treatment |  |  | 0.3 |
| None | 2 (33%) | 6 (11%) |  |
| Single | 2 (33%) | 16 (30%) |  |
| Combined | 2 (33%) | 31 (58%) |  |
| Median follow-up (months) | 3 (0, 8) | 15 (6, 30) | 0.045 |

<sup>7</sup> Statistics presented: n (%); median (IQR)
<sup>2</sup> Statistical tests performed: Fisher's exact test; chi-square test of independence; Wilcoxon rank-sum test

**Table 3.** Multivariable Cox regression survival analysis from pooled data of 59 PCNSL patients with known MYD88 mutation status and treatment information from Scottish cohort (N=41) and Yamada *et al*. 2015 ^9^ (N=18).

| Characteristic | HR <sup>7</sup> | 95% CI <sup>7</sup> | p-value |
| --- | --- | --- | --- |
| **MYD88 L265P** |  |  |  |
| Wild type | — | — |  |
| Mutated | 0.31 | 0.13, 0.77 | **0.012** |
| **Age** | 0.81 | 0.54, 1.23 | 0.3 |
| **Treatment** |  |  |  |
| None | — | — |  |
| Single | 0.17 | 0.06, 0.46 | **<0.001** |
| Combined | 0.07 | 0.02, 0.18 | **<0.001** |
| <sup>7</sup> HR = Hazard Ratio, CI = Confidence Interval |  |  |  |

### Survival analysis

Survival function estimated by Kaplan-Meier method stratified by *MYD88*, sex, age, treatment, tumour location and tumour extent are presented in Figure 1. Apart from treatment, none of these parameters were associated with better overall survival. As expected, treated patients had a significantly increased overall survival in comparison to untreated patients. The estimated median OS by Kaplan-Meier method for single and combined treatment regimens were 15 (95%CI: 4-40) and 30 (95%CI: 16-56) months, respectively. Median OS for untreated patients was 1 month. The pooled analysis revealed similar findings for age, sex and treatment (Supplementary Figure S1). There was a significant advantage in survival for *MYD88* mutated PCNSL cases, but the numbers were low (only 6 wild type *MYD88* patients).

**Figure 1.**
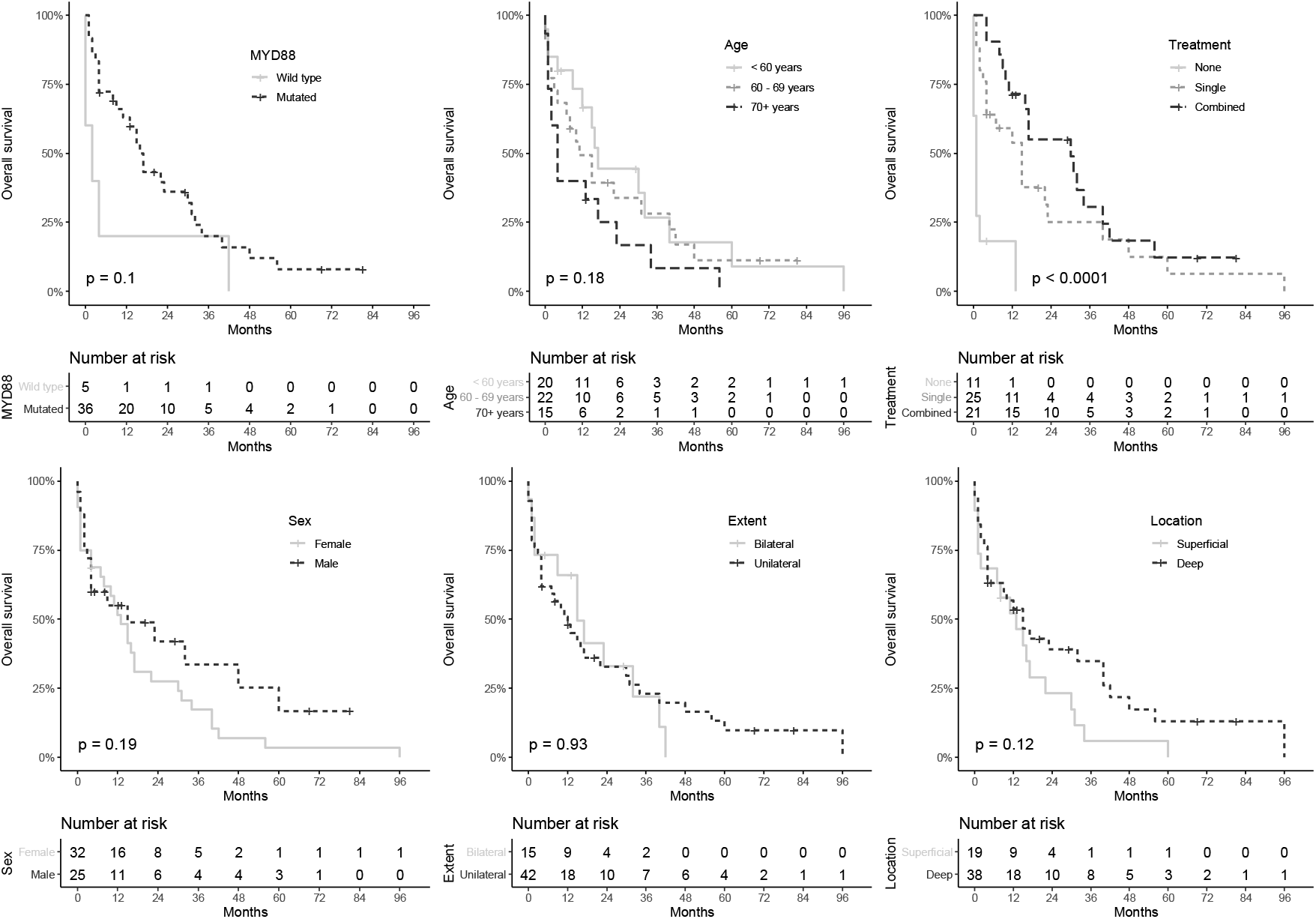
Kaplan-Meier estimation of the overall survival for 57 Scottish PCNSL patients. Overall survival was defined as the time from the date of surgery to the date of death or with censoring on the date of last available follow up.

Multivariate analysis of Scottish cohort (n=41) revealed two significant findings (Figure 2 and Supplementary Table S3). Firstly, it was confirmed that single (n=14) (HR 0.194, 95%CI: 0.07-0.55, p=0.002) and combined (n=19) (HR 0.079, 95%CI: 0.026-0.25, p<0.001) treatments were associated with survival advantage to no treatment. Secondly, mutant *MYD88* (n=36) was associated with a significant survival advantage relative to wildtype *MYD88* (n=5) (HR 0.34, 95%CI: 0.12-0.95, p=0.0389). These findings were further validated in pooled multivariate analysis cohort (n=59) (HR 0.31, 95%CI: 0.13-0.77, p=0.012) (Figure 3 and Supplementary Table S4).

**Figure 2.**
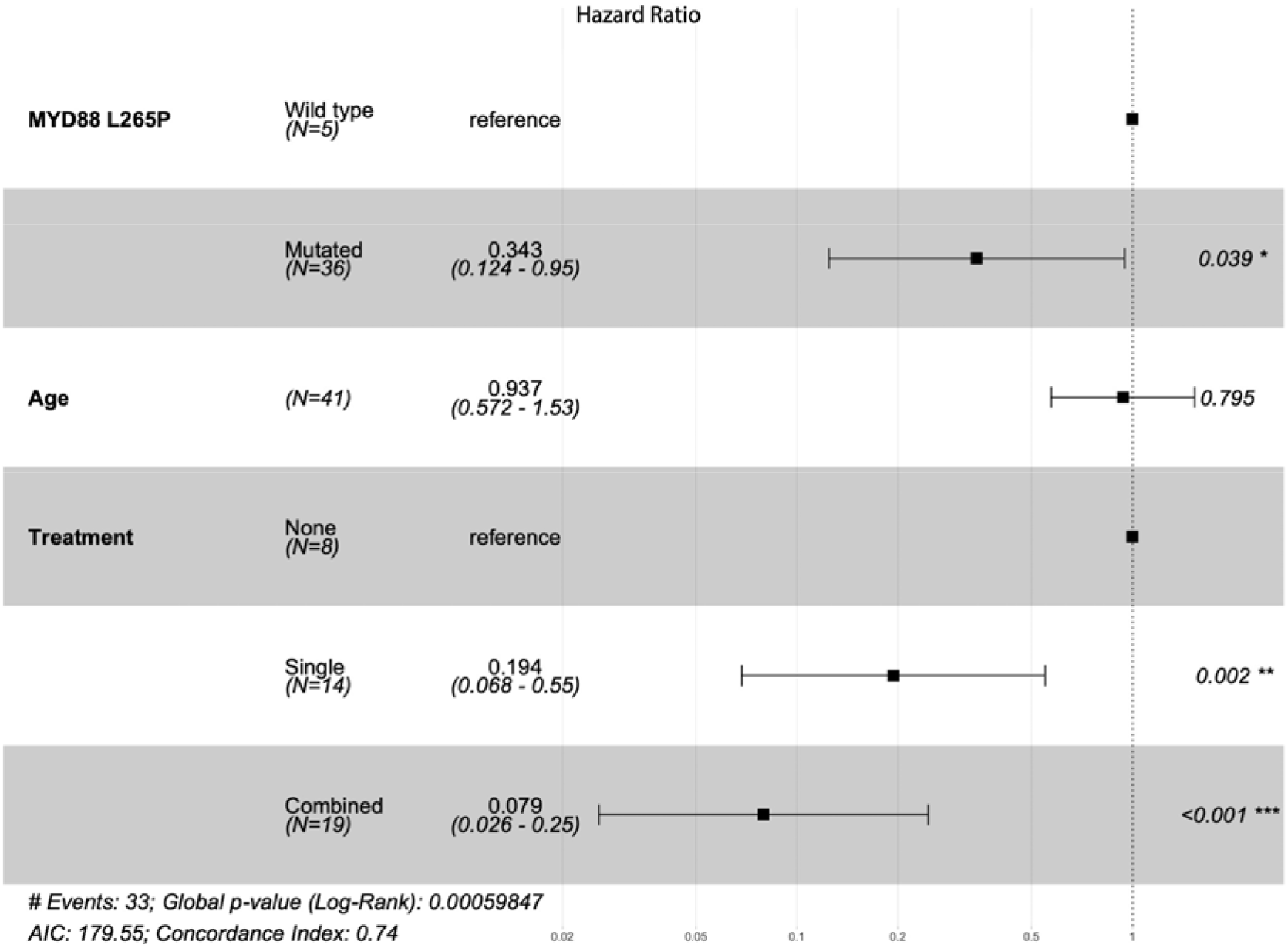
Forest plot for Cox proportional hazards model of multivariate analysis for 41 Scottish PCNSLs with known MYD88 L265P mutation status and treatment showing hazard ratios, 95% confidence intervals and p-values.

**Figure 3.**
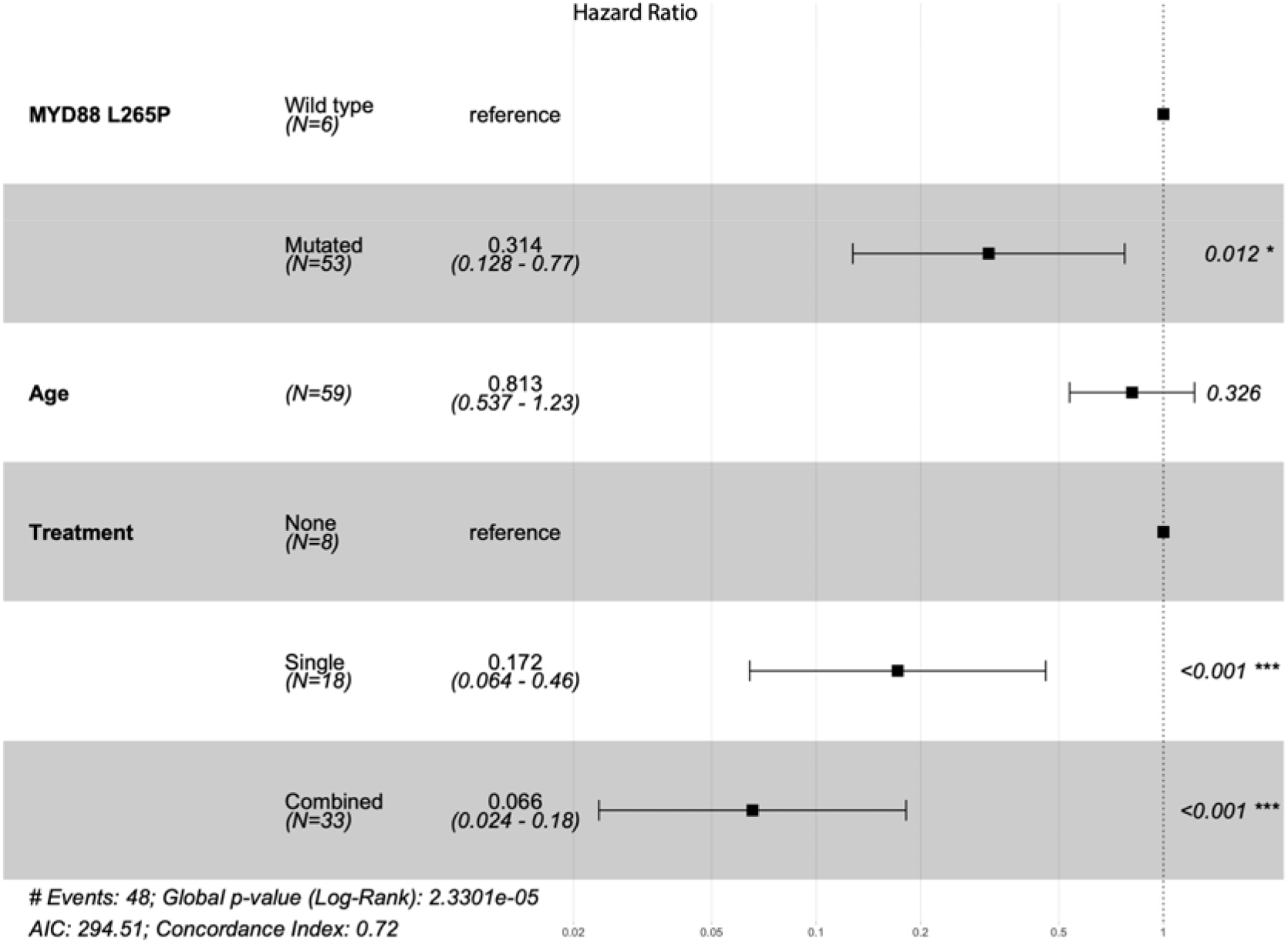
Forest plot for Cox proportional hazards model of multivariate analysis from pooled data of 59 PCNSLs with known MYD88 L265P mutation status and treatment showing hazard ratios, 95% confidence intervals and p-values.

## DISCUSSION

This retrospective single-centre study describes 57 PCNSLs with emphasis on the significance of *MYD88* L265P mutation on overall survival in immunocompetent patients who received different treatment regimens. We show that *MYD88* mutation in PCNSLs offers survival advantage. Considering the increasing importance of molecular markers in prognostication and treatment strategy, our results highlight emerging clinicopathological and molecular factors for this rare patient group with PCNSL.

Consistent with previous reports ^9,13,18,27–32^, we found a high frequency of the *MYD88* L265P mutation in PCNSLs. A recent meta-analysis by Lee *et al* reported the prevalence of this mutation in 59.8% (95%CI: 42.2-75.2%) of CNS cases^10^. The higher frequency of 88% reported in our study may reflect all cases belonging to non-GCB subtype of PCNSL. This subtype is known to be more common in PCNSLs ^31^. Moreover, non-GCB lymphoid malignancies from other sites have higher rates of this particular mutation^10,13^. Non-GCB subtype is characterised by constitutive activation of NF-κB (nuclear factor kappa-light-chain enhancer of activated B cells) pathway ^33^, while *MYD88* mutation is an oncogenic driver of NF-κB pathway ^30^. It remains uncertain, however, as to why CNS lymphomas have an increased incidence of dysregulated NF-κB signalling at a site, the CNS, which is also considered to be immune-privileged ^30,34^. This may explain why other mutations, in addition to the *MYD88* L265P, are necessary to co-exist in order to have a prognostic impact in these rare lymphomas ^8,29,34^.

In contrast to the previous studies, we demonstrate that the presence of mutant *MYD88* is associated with better survival in PCNSL patients. The prognostic value of *MYD88* mutation in systemic DLBCLs has been debated for some time with studies providing data for both sides of the argument ^10^. For PCNSLs several studies reported no effect on overall survival ^9,11,19,20^ with only two studies reporting an unfavourable outcome ^21,22^. There are, nevertheless, important methodological differences between ours and the previous reports making direct comparisons between the studies difficult. Discrepancies may be due to different analytic approaches. Association between molecular markers and survival is often confounded by clinical variables. Moreover, only three of the seven identified studies ^9,18–22,35^ performed multivariate analyses; two reporting adverse impact on OS ^21,22^ and one showing no difference ^18^. Furthermore, the discrepancies may reflect differences in biological properties of the PCNSL subtypes. Our PCNSL cases were exclusively non-GCB subtype and all but one ^21^ included studies did not report on DLBCL subtypes. Previous studies reported limited treatment variables ^18^, with some recruited only patients treated with chemotherapy ^21,22^. Selection bias is likely to contribute to the heterogeneous findings on the association between *MYD88* mutation and survival. Lastly, there may also be genetic differences between the different patient cohorts with the three studies originating from Japan.

Interestingly, our data suggest that among treated PCNSL cases, a combined-modality therapy may be better at prolonging survival. A recent study of PCNSLs reported that a significantly prolonged survival could be achieved with a combined-modality therapy, although unlike in our study, autologous stem cell therapy was excluded^36^. Current treatment regimens for PCNSL remain ineffective partly because CNS is inaccessible to many drugs. This may explain the lack of significant differences in overall survival between generic treatment options, such as chemotherapy. Combined-modality therapies may prolong survival but tend to lead to significant neurotoxicity. Newer treatments designed to reflect molecular tumour background may be more effective if target specific group of patients. In fact, therapeutic agents targeting *MYD88* mutations have already been tested in clinical settings. Ibrutinib, a Bruton tyrosine kinase inhibitor, inhibits NF-κB signalling pathway, and has been used for treatments of systemic non-GCB DLBCLs ^14^ and PCNSLs ^16,37^. An 83% partial response rate was reported in a recent clinical trial of ibrutinib in PCNSL ^16^. Our findings suggest that identification of the *MYD88* L265P mutation in PCNSLs and the use of targeted therapy may be of benefit to such patients.

### Strengths and limitations

This study included the largest cohort of patients with PCNSL with clinical, molecular and treatment variables available. We were able to investigate the effect of *MYD88* mutation on survival accounting for major confounders. Results from the pooled cohort from the literature also supported our observation. However, investigating a rare type of tumour inevitably has issues of power. We only included age and treatment in the multivariate analysis because these are the most important confounders to *MYD88* mutation. This also avoided overfitting of the data. While it would be of interest to investigate other prognostic factors, we were unable to do so. The existing literature on PCNSL is heterogeneous and pooling of individual patient data was challenging. We only included patients with treatment information since treatment is a strong predictor of survival. Though this did not allow a larger cohort to be established, our pooled analyses included only patients with adequate clinical features to be informative. On treatment for PCNSL, there is not one standard of care and insights into efficacy of different treatment would be helpful. This study was unable to address that.

In summary, in line with previous reports, this study shows that *MYD88* L265P mutation is common in PCNSLs and it is not a prognostic factor in univariate analysis. This study reports that the presence of *MYD88* L265P mutation is associated with longer survival in PCNSL patients. While further validation of this association is warranted, our findings suggest that identification of *MYD88* mutation can identify patients who may benefit from novel targeted therapies and enhance survival.

## Data Availability

The authors confirm that all data underlying the findings are available and will be shared with the research community upon request.

## Acknowledgements

Part of this work was presented at the 120^th^ meeting of the British Neuropathological Society, 4-6 March 2019, London, UK by OEC.

## Conflict of interests

The authors declare no conflict of interest.

## Funding

MTCP is supported by Cancer Research UK Brain Tumour of Excellence Award (C157/A27589).

## Authorship statement

OEC - study design, data acquisition, analysis and interpretation, manuscript preparation; MTCP – data analysis and interpretation, manuscript preparation; LG - MYD88 analysis; AT and CS – pathology reporting; WAQ – concept and study design, data acquisition, pathology reporting. All authors reviewed the final manuscript.

## Notes

### Competing Interest Statement

The authors have declared no competing interest.

### Author Declarations

The ethical approval for this study was waived by the Tissue Governance committee of the South East Scotland SAHSC BioResource.

## REEFERENCES

1. Berglund M, Thunberg U, Amini RM, et al. Evaluation of immunophenotype in diffuse large B-cell lymphoma and its impact on prognosis. Mod Pathol. Published online 2005.

2. Swerdlow SH, Campo E, Harris NL, et al. WHO Classification of Tumours of Haematopoietic and Lymphoid Tissues. Lyon, France. World Heal Organ Calssification Tumours Haematop Lymphoid Tissue. Published online 2017.

3. Hans CP, Weisenburger DD, Greiner TC, et al. Confirmation of the molecular classification of diffuse large B-cell lymphoma by immunohistochemistry using a tissue microarray. Blood. 2004;103(1):275–282.

4. Deckert M, Brunn A, Montesinos-Rongen M, Terreni MR, Ponzoni M. Primary lymphoma of the central nervous system-a diagnostic challenge. Hematol Oncol. 2014;32(2):57–67.

5. Fox CP, Phillips EH, Smith J, et al. Guidelines for the diagnosis and management of primary central nervous system diffuse large B-cell lymphoma.(Report). Br J Haematol. 2019;184(3):348.

6. Carnevale J, Rubenstein JL. The Challenge of Primary Central Nervous System Lymphoma. Hematol Oncol Clin North Am. 2016;30(6):1293–1316.

7. Compagno M, Lim WK, Grunn A, et al. Mutations of multiple genes cause deregulation of NF-kappaB in diffuse large B-cell lymphoma. Nature. 2009;459(7247):717–721.

8. Poulain S, Boyle EM, Tricot S, et al. Absence of CXCR4 mutations but high incidence of double mutant in CD79A/B and MYD88 in primary central nervous system lymphoma. Br J Haematol. 2015;170(2):285–287.

9. Yamada S, Ishida Y, Matsuno A, Yamazaki K. Primary diffuse large B-cell lymphomas of central nervous system exhibit remarkably high prevalence of oncogenic MYD88 and CD79B mutations. Leuk Lymphoma. 2015;56(7):2141–2145.

10. Lee J-H, Jeong H, Choi J-W, Oh H, Kim Y-S. Clinicopathologic significance of MYD88 L265P mutation in diffuse large B-cell lymphoma: a meta-analysis. Sci Rep. 2017;7(1):1785.

11. Nayyar N, White MD, Gill CM, et al. MYD88 L265P mutation and CDKN2A loss are early mutational events in primary central nervous system diffuse large B-cell lymphomas. Blood Adv. Published online 2019.

12. Zorofchian S, Lu G, Zhu JJ, et al. Detection of the MYD88p.L265P mutation in the CSF of a patient with secondary central nervous system lymphoma. Front Oncol. Published online 2018.

13. Ngo VN, Young RM, Schmitz R, et al. Oncogenically active MYD88 mutations in human lymphoma. Nature. 2011;470(7332):115–119.

14. Wilson WH, Young RM, Schmitz R, et al. Targeting B cell receptor signaling with ibrutinib in diffuse large B cell lymphoma. Nat Med. 2015;21(8):922–926.

15. Bernard S, Goldwirt L, Amorim S, et al. Activity of ibrutinib in mantle cell lymphoma patients with central nervous system relapse. Blood. 2015;126(14):1695–1698.

16. Lionakis MS, Dunleavy K, Roschewski M, et al. Inhibition of B Cell Receptor Signaling by Ibrutinib in Primary CNS Lymphoma. Cancer Cell. 2017;31(6):833–843.e5.

17. Chamoun K, Choquet S, Boyle E, et al. Ibrutinib monotherapy in relapsed/refractory CNS lymphoma: A retrospective case series. Neurology. 2017;88(1):101–102.

18. Nakamura T, Tateishi K, Niwa T, et al. Recurrent mutations of CD79B and MYD88 are the hallmark of primary central nervous system lymphomas. Neuropathol Appl Neurobiol. 2016;42(3):279–290.

19. Zhou Y, Liu W, Xu Z, et al. Analysis of Genomic Alteration in Primary Central Nervous System Lymphoma and the Expression of Some Related Genes. Neoplasia. 2018;20(10):1059–1069.

20. Zheng M, Perry AM, Bierman P, et al. Frequency of MYD88 and CD79B mutations, and MGMT methylation in primary central nervous system diffuse large B-cell lymphoma. Neuropathology. 2017;37(6):509–516.

21. Hattori K, Sakata-Yanagimoto M, Okoshi Y, et al. MYD88 (L265P) mutation is associated with an unfavourable outcome of primary central nervous system lymphoma. Br J Haematol. 2017;177(3):492–494.

22. Takano S, Hattori K, Ishikawa E, et al. MyD88 Mutation in Elderly Predicts Poor Prognosis in Primary Central Nervous System Lymphoma: Multi-Institutional Analysis. World Neurosurg. 2018;112:e69–e73.

23. Vinet L, Zhedanov A. A ‘missing’ family of classical orthogonal polynomials. J Phys A Math Theor. 2011;44(8):085201.

24. Choi WWL, Weisenburger DD, Greiner TC, et al. A New Immunostain Algorithm Classifies Diffuse Large B-Cell Lymphoma into Molecular Subtypes with High Accuracy. Clin Cancer Res. 2009;15(17):5494–5502.

25. Jiménez C, Del Carmen Chillón M, Balanzategui A, et al. Detection of MYD88 L265P mutation by real-time allele-specific oligonucleotide polymerase chain reaction. Appl Immunohistochem Mol Morphol. Published online 2014.

26. King RL, Goodlad JR, Calaminici M, et al. Lymphomas arising in immune-privileged sites: insights into biology, diagnosis, and pathogenesis. Virchows Arch. Published online 2019.

27. Montesinos-Rongen M, Godlewska E, Brunn A, Wiestler OD, Siebert R, Deckert M. Activating L265P mutations of the MYD88 gene are common in primary central nervous system lymphoma. Acta Neuropathol. Published online 2011.

28. Yu X, Li W, Deng Q, et al. *MYD88* L265P Mutation in Lymphoid Malignancies. Cancer Res. 2018;78(10):2457 LP - 2462.

29. Ou A, Sumrall A, Phuphanich S, et al. Primary CNS lymphoma commonly expresses immune response biomarkers. Neuro-oncology Adv. 2020;2(1):vdaa018.

30. Braggio E, Van Wier S, Ojha J, et al. Genome-Wide Analysis Uncovers Novel Recurrent Alterations in Primary Central Nervous System Lymphomas. Clin Cancer Res. 2015;21(17):3986.

31. Camilleri-Broet S, Crinière E, Broet P, et al. A uniform activated B-cell–like immunophenotype might explain the poor prognosis of primary central nervous system lymphomas: analysis of 83 cases. Blood. 2006;107(1):190–196.

32. Vater I, Montesinos-Rongen M, Schlesner M, et al. The mutational pattern of primary lymphoma of the central nervous system determined by whole-exome sequencing. Leukemia. 2015;29(3):677–685.

33. Lim K-H, Yang Y, Staudt LM. Pathogenetic importance and therapeutic implications of NF-[kappa]B in lymphoid malignancies.(Report). Immunol Rev. 246:359.

34. Kraan W, Horlings HM, van Keimpema M, et al. High prevalence of oncogenic MYD88 and CD79B mutations in diffuse large B-cell lymphomas presenting at immune-privileged sites. Blood Cancer J. 2013;3(9):e139–e139.

35. Gonzalez-Aguilar A, Idbaih A, Boisselier B, et al. Recurrent mutations of MYD88 and TBL1XR1 in primary central nervous system lymphomas. Clin Cancer Res. 2012;18(19):5203.

36. Niparuck P, Boonsakan P, Sutthippingkiat T, et al. Treatment outcome and prognostic factors in PCNSL. Diagn Pathol. 2019;14(1):56.

37. Grommes C, Pastore A, Palaskas N, et al. Ibrutinib unmasks critical role of bruton tyrosine kinase in primary CNS lymphoma. Cancer Discov. Published online 2017.

